# Using real-time data to guide decision-making during an influenza pandemic: a modelling analysis

**DOI:** 10.1101/2021.06.09.21258618

**Authors:** David J. Haw, Matthew Biggerstaff, Pragati Prasad, Joseph Walker, Bryan Grenfell, Nimalan Arinaminpathy

## Abstract

Influenza pandemics typically occur in multiple waves of infection, often associated with initial emergence of a novel virus, followed (in temperate regions) by a later resurgence accompanying the onset of the annual influenza season. Here, we examined whether data collected from an initial pandemic wave could be informative, for the need to implement non-pharmaceutical measures in any resurgent wave. Drawing from the 2009 H1N1 pandemic in 10 states in the USA, we calibrated simple mathematical models of influenza transmission dynamics to data for virologically confirmed hospitalisations during the initial ‘spring’ wave. We then projected pandemic outcomes (cumulative hospitalisations) during the fall wave, and compared these projections with data. Model results show reasonable agreement for all states that reported a substantial number of cases in the spring wave. Using this model we propose a probabilistic decision framework that can be used to determine the need for pre-emptive measures such as postponing school openings, in advance of a fall wave. This work illustrates how model-based evidence synthesis, in real-time during an early pandemic wave, could be used to inform timely decisions for pandemic response.

## Introduction

Even as the current coronavirus pandemic causes morbidity, mortality and societal disruption on a global scale, the threat of similar disruptions from pandemic influenza continues unabated [10]. The 1918 H1N1 pandemic caused an estimated 50 million deaths worldwide, with a similar case fatality rate to COVID-19 [1,3]. Today, highly pathogenic strains of avian influenza continue to cause sporadic infections in humans, with occasional instances of human-to-human transmission [13]: preparedness for a future influenza pandemic thus remains as important as ever.

A marked feature of influenza epidemiology is its pronounced seasonality, with already-established influenza viruses causing epidemics each winter in temperate regions of the world. Such seasonality is likely to arise from a combination of factors, and has been associated with environmental conditions including absolute humidity [19], as well as increased transmission amongst schoolchildren with the post-holiday opening of school terms [7,12]. These seasonal drivers have played a strong role in shaping the dynamics of pandemic, as well as seasonal, influenza. For example, in the USA in 2009, the novel H1N1 virus caused a ‘spring wave’ from April to July, during which an estimated 1.8m-5.7m people experienced symptomatic infection and 9,000-21,000 were hospitalised [17]. The subsequent onset of the influenza season in October 2009 was accompanied by a strong resurgence of the virus (the ‘fall wave’), resulting an estimated 60.8m total infections and 274,304 hospitalisations by April 2010 [20]. Similar multi-wave behaviour emerged in other temperate countries, with the UK experiencing three successive waves [16,6]. If novel influenza viruses can emerge at any time of year in temperate countries, it is more likely than not that they would do so outside the normal influenza season, which typically spans a 4 month period (late October to late February) in the Northern Hemisphere.

For any future influenza pandemic, population surveillance data collected from any initial, out-of-season wave could therefore give important information for characterising severity and transmissibility. Could such data be used to predict the potential health impact of a subsequent fall/winter wave? If so, could such projections be used in real time, to trigger pre-emptive control measures in advance of that wave? In this work we addressed these questions using a mathematical model of influenza transmission dynamics, with a focus on pandemic spread in the USA. We present examples of how this framework can be used to guide real-time decision-making for future pandemics.

## Results

Figure 1 shows FluSurv-NET data for virologically confirmed hospitalisations with pH1N1 during the spring and fall waves of the 2009 pandemic, for 10 states in the USA in which this data was available. The figure illustrates the difference in size between the two waves. Notably, the geographically southern states of Georgia, New Mexico and Tennessee reported only sparse data in the spring/summer period: these same states also showed substantially fewer cases being reported in the fall wave per capita, when compared to other states.

**Figure 1:**
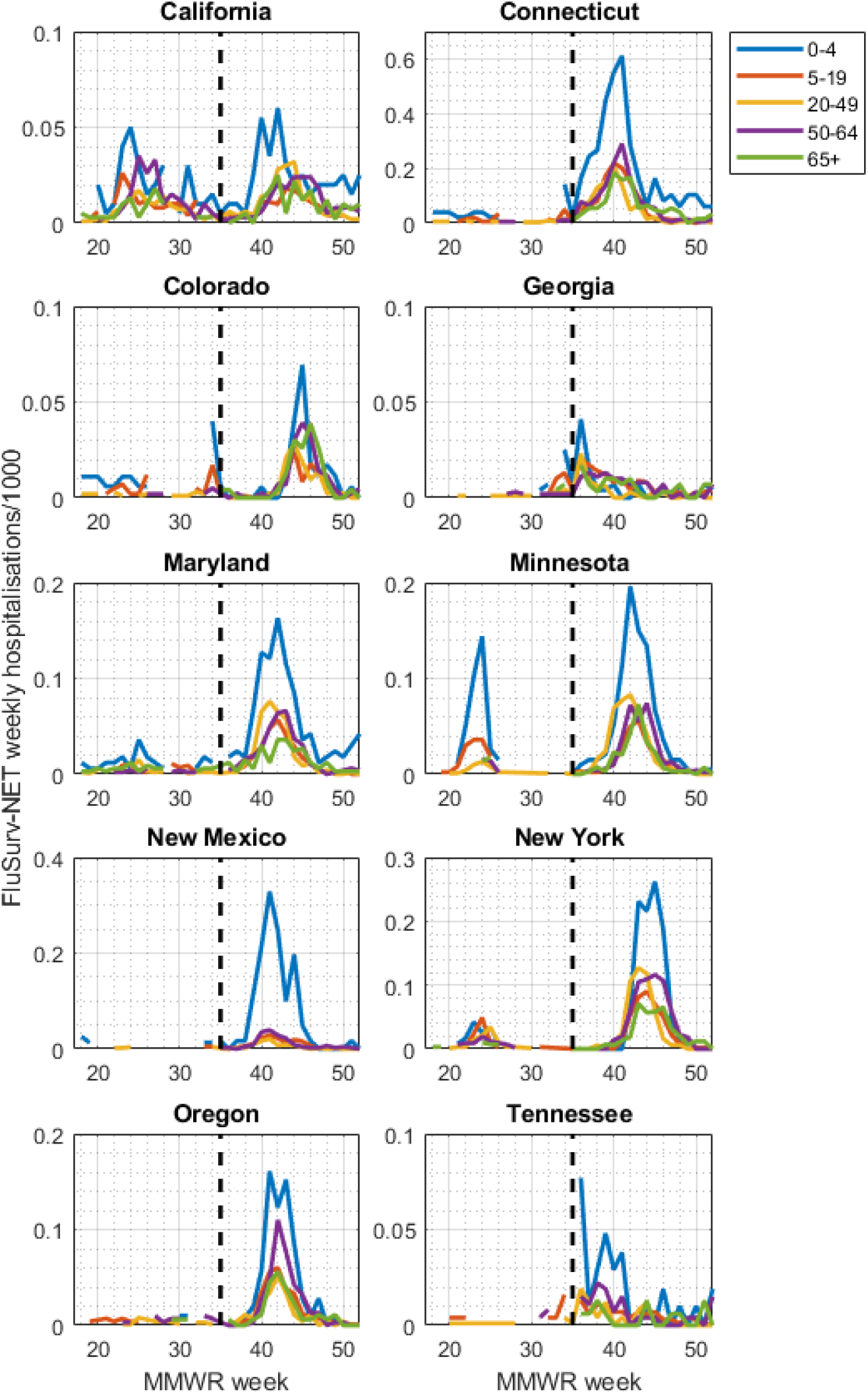
FluSurv-NET data from the 2009 H1N1 pandemic in the USA. Shown is weekly, age-specific data collected by the US Centers for Disease Control and Prevention (CDC), for hospitalisations that were virologically confirmed as being pandemic H1N1. Each colour denotes a different age group, as indicated by the legend. Panels show data from the different states reporting FluSurv-NET data. Weeks are numbers along the x-axis according to MMWR numbering with, for example, week 35 corresponding to the week beginning on Sunday 30th August. Note that the *y*-axis varies between states. As described in the main text, several of these states (e.g. California) show clear signs of distinct spring and fall waves. For the purpose of the model, we used these data in combination with CDC estimates for the proportion of symptomatic cases that are hospitalised, virologically tested, and reported through this dataset.

To model these data, we calibrated a deterministic, compartmental model of influenza transmission dynamics, with five age groups: <4 years old (yo), 5-19yo, 20-49yo, 50-64yo, and >65yo (see Materials and Methods). We calibrated this model to data from the spring wave illustrated in Figure 1, i.e. from months April-August. For each location we then used the calibrated model to project the fall wave dynamics. Figure 2 shows results in the example of California, illustrating both spring wave calibration, and fall wave projections. Figure S2 shows outputs disaggregated by state, and Figure S3 illustrates the performance of the Bayesian Markov Chain Monte Carlo used to perform these calibrations. Although the model tends to estimate fall wave peak timing somewhat earlier than actually occurred in the data (week 41 vs week 42 for aggregate results, week 40 vs week 44 for Connecticut), there is reasonable agreement for the size of the fall wave (comparing areas under the curve for model projections and data for incidence). For the remainder of this analysis, we focus on the cumulative burden in the fall wave rather than peak timing.

**Figure 2:**
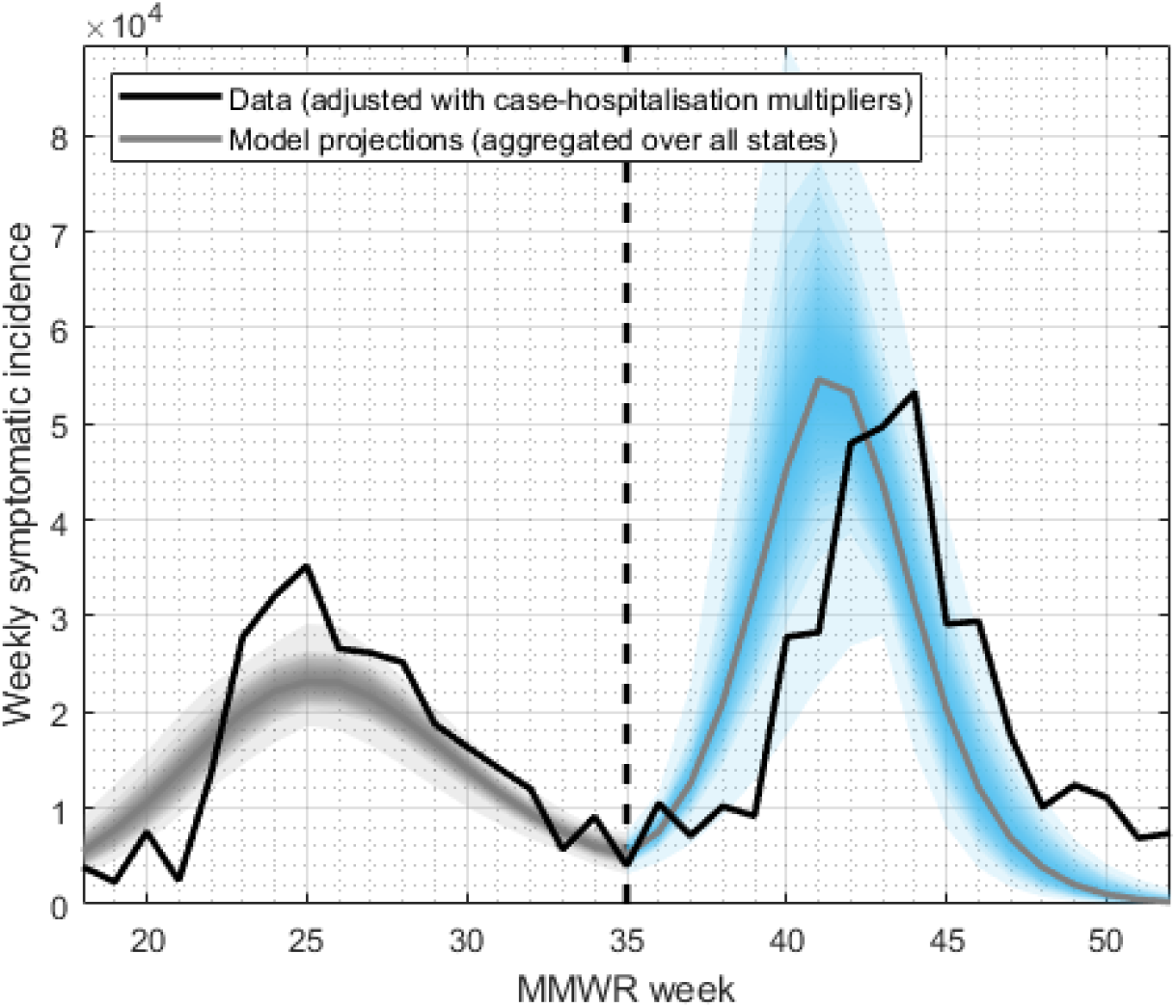
Illustration of the modelling approach, and of model projections, in the example of California. For each state shown in Fig.1, we calibrated the model to the epidemic data from the spring wave (black line, to the left of the vertical dashed line, with aggregated model projections shown in grey shaded area). Using this calibrated model, we projected simulations forward into the fall, taking account of the effect of school openings and environmental forcing (blue shaded area). Although the model projection for epidemic peak timing varied in accuracy across states, our subsequent analysis concentrates on cumulative burden (area under the curve). See Figure S1 for results for other states.

Figure 3 shows a state-wise disaggregation for the cumulative projected hospitalisations in the fall wave. Model projections show good agreement in 7 of the 10 states studied. However, model projections appear less accurate in Georgia, New Mexico and Tennessee, where the model substantially overestimates cumulative hospitalisations in the fall wave. As noted above, these are the states with only sparse data reported in both spring and fall waves.

**Figure 3:**
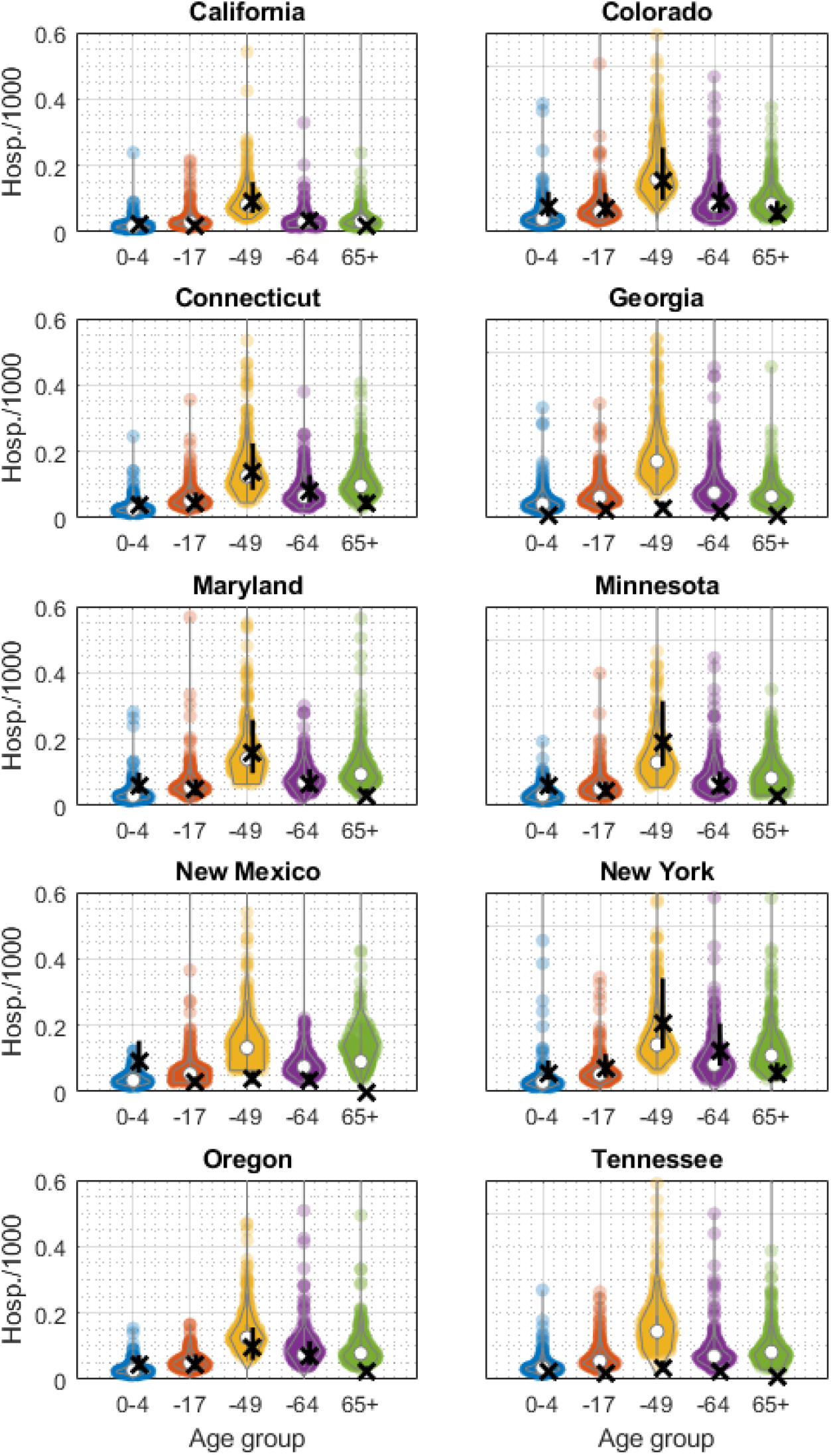
Model projections for cumulative hospitalisations in the fall wave. Each panel shows a different state. Crosses in black show data, while coloured points show model-based projections, with each point representing the result of a single sample from the posterior density.

For those states showing reasonable model performance, we next examined how this framework could be operationalised, to trigger pre-emptive interventions in advance of the fall wave. Such interventions could involve, for example, physical distancing orders; pre-emptive school closures; or other non-pharmaceutical measures aimed at reducing opportunities for transmission. As such measures are typically costly and disruptive, any decision to implement them must carefully balance these disruptions against the risks of widespread morbidity and mortality.

As an illustrative example in the current analysis, we concentrated on pre-emptive school closures (i.e. postponing the start of the school term), until a vaccine becomes available. To inform our assumptions for vaccine roll-out, we assumed the same trajectory as in the H1N1 pandemic, when a vaccination programme was in October, ultimately to cover over 25% of the population (see Figure S4). In a hypothetical scenario where pre-emptive school closures are implemented for a 2009-like pandemic, Figure 4 illustrates the reduction in fall wave burden, as a function of different durations of intervention. The figure illustrates, for example, that a 10-week postponement in school opening, together with the impact of the vaccine rollout, could reduce cumulative hospitalisations in the fall wave by 72% (95% CrI 38-90%).

**Figure 4:**
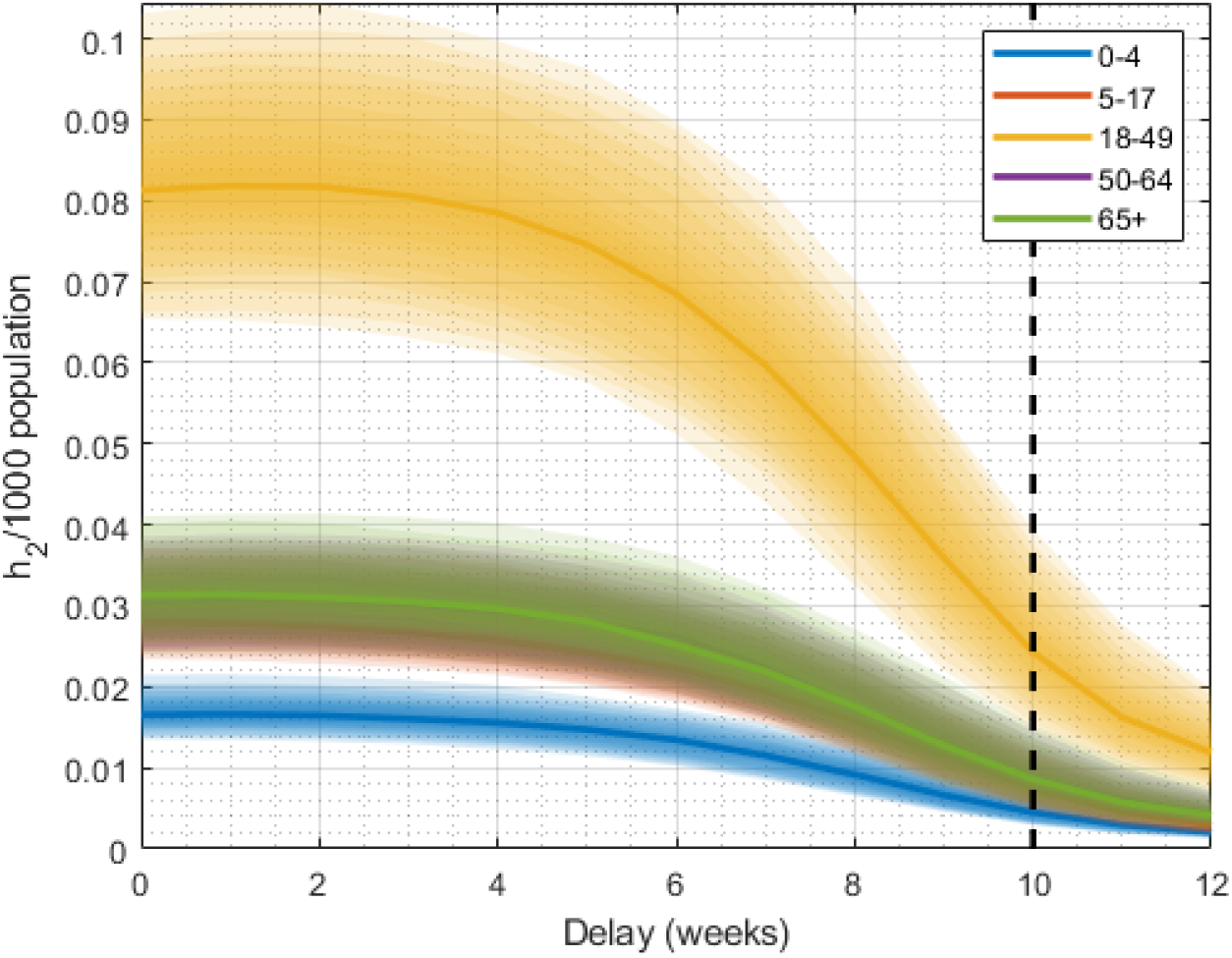
Projected hospitalisations (California) with delayed school openings, assuming the same efficacy and timing of vaccine rollout as occurred in 2009. Each colour shows a different age group as indicated by the legend, while shaded areas show 25-75th percentiles, with 2009 vaccination coverage/efficacy. The vertical dashed line represents the candidate delay of 10 weeks usedto illustrate our decision framework in figure 5.

As a decision tool for when to trigger such measures, we defined the ‘probabilistic risk score’ (PRS) as the probability that cumulative hospitalisations in the fall wave will exceed a threshold of *h* per capita. We assumed that this risk score would be evaluated at the end of the spring wave, and that pre-emptive school closures would be triggered if PRS exceeds a given threshold *p*. In practice, both *h* and *p* would be determined by a policymaker, and can be interpreted as reflecting considerations of healthcare capacity (cumulative hospitalisations, *h*), alongside tolerance of uncertainty (*p*). As an illustrative example, we assumed a scenario where *h* = 1, 500 and *P* = 0.1. Figure 5 illustrates this decision tool being applied to the H1N1 pandemic in California, as well as an alternative scenario with a hypothetical virus having twice the severity (i.e. the same infectivity, but twice the risk of hospitalisation). Under these parameters, a 2009-like virus would not trigger the intervention (blue line), whereas a more severe virus would do so (solid red line). The FluSurv-NET data set estimates 1, 013 hospitalisations in California during the second wave, consistent with our decision not to trigger an intervention. Moreover in this second scenario, pre-emptive school closures for 10 weeks would bring PRS substantially below the threshold (dashed red line).

**Figure 5:**
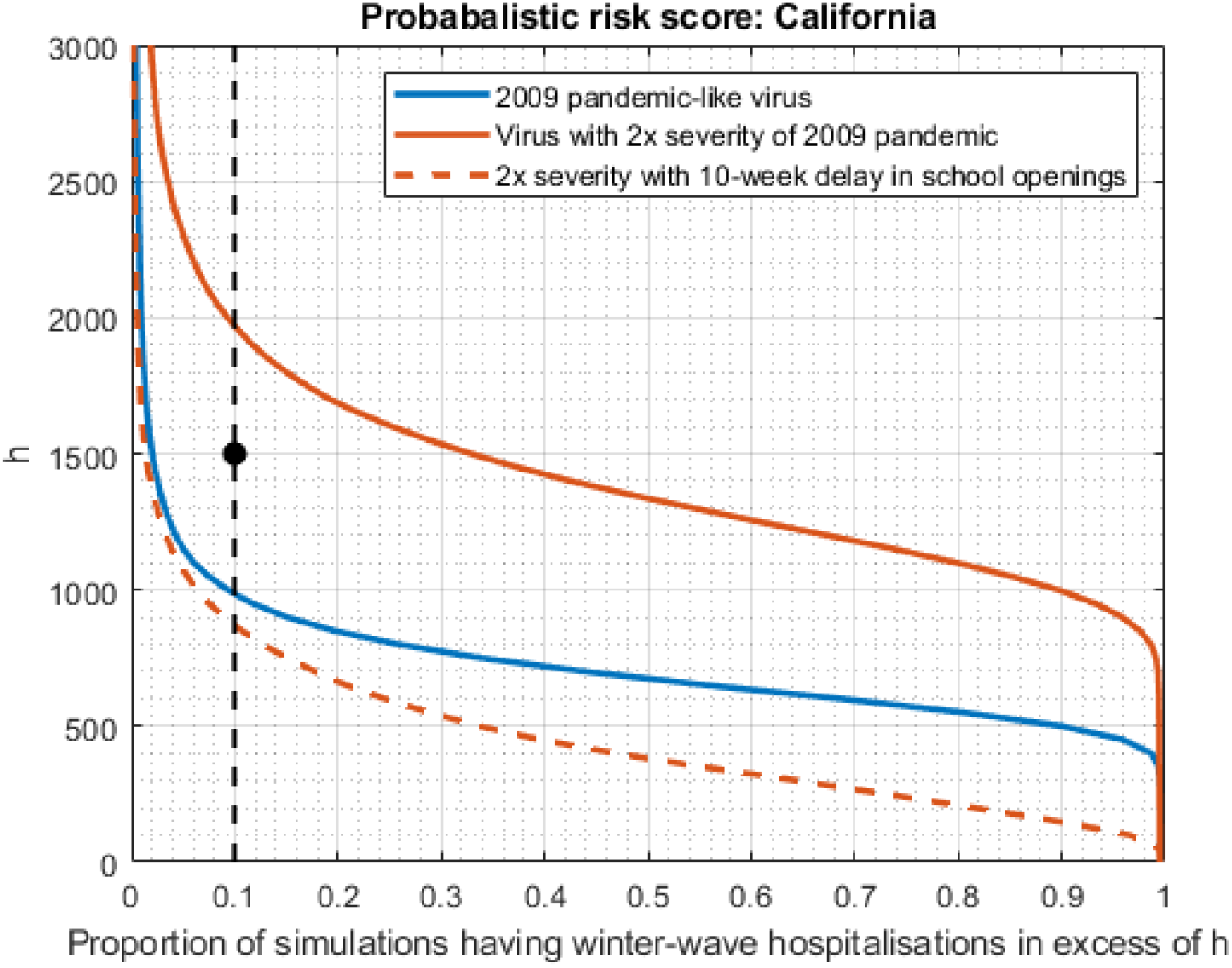
Proposed decision framework for triggering pre-emptive non-pharmaceutical interventions (NPIs), in advance of the fall wave. Shown, for illustration, is the example of California. These plots can be interpreted as cumulative probability distributions, for the total hospitalisations projected in the fall wave. As described in the main text, we define a ‘probabilistic risk score’ (PRS) as the probability that fall wave hospitalisations will increase a given threshold, *h*. We assume that pre-emptive interventions would be triggered if PRS exceeds some threshold probability *P*, with both H and P determined by a policymaker. The figure shows an illustrative scenario where *h* = 1, 500 cumulative hospitalisations, and *P* = 0.1 (‘reference point’, shown as a black dot). Any model-based projections can be represented as a downward-sloping curve on this plot: pre-emptive interventions would be triggered if the curve intersects the vertical, dashed line at any point above the reference point. As examples, the blue curve shows model projections for a 2009-pandemic-like virus in California (i.e. corresponding to Fig.3A), a scenario that would not trigger pre-emptive interventions. The solid red curve shows an alternative scenario, of a virus that is equally infectious, but twice as severe (i.e. having twice the risk of hospitalisation given infection). Such a virus would trigger pre-emptive interventions; the dashed red curve shows the reduction in hospitalisation risk that would occur, in a scenario where school opening is postponed for 10 weeks until vaccine rollout is underway (assuming the same vaccine introduction and rollout scenario as occurred in 2009-2010, in response to the pandemic).

## Discussion

In any future influenza pandemic, early and accurate information will be critical in deciding how best to respond. Here we have examined how mathematical modelling of transmission dynamics could be used to analyse surveillance data in the early stages of a pandemic, to inform decisions for pre-emptive non-pharmaceutical interventions, in advance of any second wave of infection. Notably, although our approach is based on a relatively simple, compartmental model of influenza transmission, for states where there is sufficient data, this approach shows reasonable projections for cumulative fall wave burden (Fig.3). A key benefit of such a modelling approach is that it can be readily deployed in real time: we illustrate how this model could be applied in a probabilistic way, to inform decisions for pre-emptive school closures (Fig.4).

Model calibration to an unmitigated phase allows us to capture important epidemiological properties of an outbreak, most crucially the basic reproductive number. Though such calibration does not require an entire wave to be unmitigated, any changes in contact patterns during the calibration period is likely to bring additional uncertainty into the model.

Our modelling approach performs less well in states having only sparse data for the spring wave (Figures 1 and 3). Such sparseness could be explained either by under-reporting, or by a genuinely lower level of influenza activity in these locations. However, it is notable that these states also reported systematically low numbers in the fall wave as well (Figures 3 and S2), suggesting that data reporting may be the driving factor. We focused on data for virologically confirmed hospitalisation because it was the least affected by changes in testing practices, during the pandemic [18]. Nonetheless, an important area for future work is to explore the potential for incorporating other forms of data as well, including syndromic and virological surveillance collected from the primary care level and above [14]. Combining different streams of data in this way could offer a helpful approach, to compensate for shortfalls in any individual data stream.

As an example of non-pharmaceutical interventions, we have modelled pre-emptive school closures, i.e. postponing the opening of schools. We note that our estimates for the impact of these measures are driven by modelled variations in the age-specific contact matrix, depending on whether schools are in or out of session. In turn, these variations are derived from estimated contact rates in an education setting[2]. In future, such estimates would benefit from primary evidence for the ‘real world’ impact that could arise from pre-emptive school closures. Because much of the available, primary evidence arises from closures that occurred during the course of an epidemic, it is likely to underestimate the impact of pre-emptive measures. The absence of influenza cases in 2020-2021 is indicative that the combination of non-pharmaceutical interventions employed to mitigate he spread of COVID-19 prevented major influenza outbreaks, though school closures were only a part of this effort [21]. Available evidence is equivocal for COVID-19, but the apparently milder natural history of infection in children, in comparison with influenza, may limit its generalizibility. Nonetheless, other statistical analysis, taking advantage of country-level variations in school opening dates, suggests that later school openings are indeed associated with reduced epidemic peaks [4].

As with any modelling study, our analysis has limitations to note. Our mathematical model involves several simplifications: averaging at the state level, it does not address the marked intra-state, spatial heterogeneity seen in the 2009 pandemic [8], indeed heterogeneity that is likely to be displayed by any future pandemic as well. Further work could seek to address these complexities by incorporating spatial structure. However, it would be important for any such approach to maintain a balance between complexity, and rapid deployability, during a pandemic. As noted above, our work illustrates that even a simple model is able to capture the cumulative fall wave burden in reasonable agreement with the data. Nonetheless, more complex models may be helpful in better capturing the peak timing of the fall wave: an estimate that could be equally important for decision-making, as the cumulative burden. Additionally, our estimates depend on case-to-hospitalisation multipliers, which were estimated during the course of the 2009 pandemic by combining careseeking interviews with other sources of evidence [9]. In any future pandemic, an application of this approach would likewise necessitate such evidence generation, which would need to be implemented rapidly, during the few months of the initial wave. Alternatively, establishing readiness for serological surveillance [5] would likewise provide critical information during the course of a future pandemic, to help translate hospitalisation and syndromic surveillance data to actual burden in the community.

In conclusion, the ongoing coronavirus emergency has highlighted the difficult choices faced by decision-makers, in the face of a pandemic: how to weigh the societal disruptions of sweeping interventions against the need for rapid, decisive action to mitigate the spread of a dangerous pathogen. While surveillance alone plays a critical role in informing these decisions, dynamical analysis of this data could offer additional, important insights for pre-emptive action. We have illustrated our methodology with respect to delayed school openings in order to mitigate a fall wave, though a broader application is feasible, in which real-time data informs parameter uncertainty in a simple model, and the forward projections under different scenarios are used to inform institutional closures or behavioural change. Overall, this analysis serves to illustrate how surveillance data can be analysed with rapid, simple models, in order inform projections on the need for future intervention.

## Methods

### The Model

We use a deterministic SIR (susceptible-infectious-removed) model of influenza transmission defined as follows:

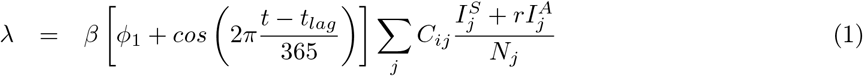

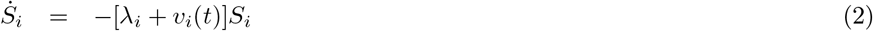

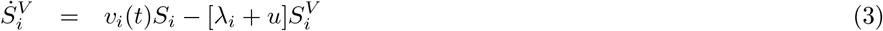

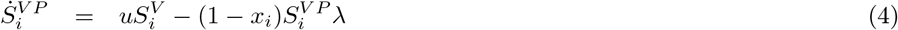

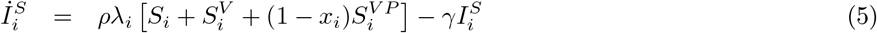

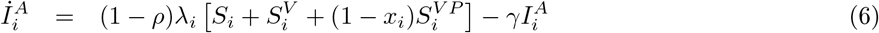

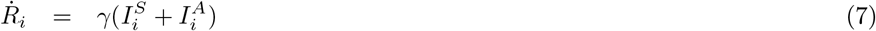

where *S*^*V*^ denotes vaccinated susceptibles, *S*^*V P*^ vaccinated susceptibles with active vaccine protection, *I*^*S*^ symptomatic infectious, *I*^*A*^ asymptomatic infectious, and the subscript *i* indexes 5 age groups (0-4, 5-19, 18-49, 50-64 and 65+). The contact matrix *C* taken from [15]. The parameter *ρ* the proportion of cases that are symptomatic, *r* the reduction in infectiousness of asymptomatic cases, *v*_*i*_(*t*) the vaccination rate in age-group *i*, and *x*_*i*_ the corresponding infection-blocking efficacy. We use the vaccination uptake and efficacy observed in 2009, which began in early October (MMWR week 39, Figure S4). We assume a mean delay of 14 days (*u* = 1*/*14) from administration of vaccine to full protection. For each FluSurv-NET location, we simulate using the corresponding age-stratified sample location populations *N*_*i*_. Hospitalisations are modelled as a multiplicative factor of symptomatic incidence, using the case-hospitalisation ratios given in [18]. Vaccination rates and hospitalisation multipliers are given at national level only.

Uncertainty in disease incidence is captured via uncertainty in case-hospitalisation multipliers. These are assumed normally distributed for each age group *i*, with mean *µ*_*i*_ and standard deviation *σ*_*i*_, given in table 1. For parameter set *θ*, our model produces simulated weekly incidence **y**(*t*), desegregate by age group. These values are divided by the corresponding hospitalisation data to yield simulated multipliers. The likelihood *L*(*θ*) is then a product of normal likelihoods *N* (*µ*_*i*_, *σ*_*i*_) over all age groups and time points.

**Table 1:**
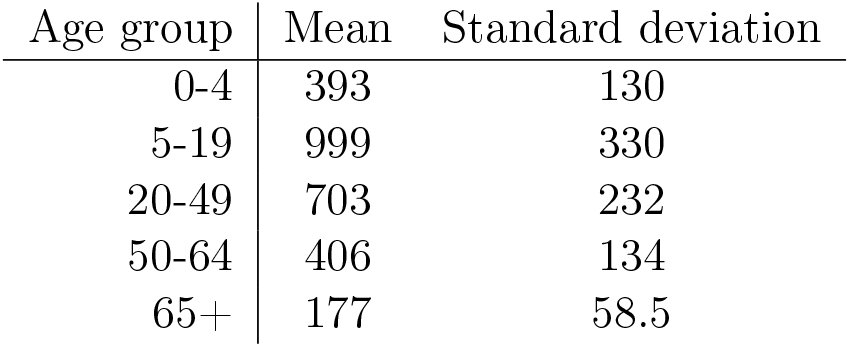
Case-hospitalisation multipliers (USA).

| Age group | Mean | Standard deviation |
| --- | --- | --- |
| 0-4 | 393 | 130 |
| 5-19 | 999 | 330 |
| 20-49 | 703 | 232 |
| 50-64 | 406 | 134 |
| 65+ | 177 | 58.5 |

We perform a Bayesian fit to the first wave only (up to 1st September, MMWR week 35) using an adaptive MCMC algorithm [11]. We allow for uncertainty in the coefficients of C and account school closures throughout this period by subtracting fixed values *δ*_1_ and *δ*_2_ from *C*_11_ and *C*_22_, determined by the education-specific contact rates given in [2]. A complete set of fitted model parameters and ranges of uniform priors is given in table S1. Sampling from the posterior density from the first wave, we project from 1st September, imposing the change in contact rates due to school openings. Together with seasonality, this generates a second wave of infection.

A delay in school openings is modelled via a delay in the change in contact rates. The probabilistic risk scores for a given state and delay in school opening are calculated by projecting the corresponding epidemic scenario for each parameter set in our posterior sample. Each simulation yields a total number of hospitalisations, which are always counted from the week of 1st September, irrespective of delay in school openings.

## Data Availability

All data (FluSurNet) is freely available online.

https://www.cdc.gov/flu/weekly/influenza-hospitalization-surveillance.htm

## Supplementary Material

**Table S1:** Model parameters and boundary values of uniform priors.

| Parameter | Min | Max |
| --- | --- | --- |
| Seasonal amplitude | 0 | 0.3 |
| Seasonal lag | -31 | 31 |
| Contact matrix (schools open) | -10% | +10% |
| Preexisting immunity | 0 | 0.4 |
| Proportion symptomatic | 0.495 (0.55-10%) | 0.605 (0.55+10%) |
| Asymptomatic infectiousness (rel.) | 0.4 | 0.6 |
| Seed time | -100 | 100 |
| Basic reproductive number | 1.305 (1.45-10%) | 1.595 (1.45+10%) |
| Infectious period (days) | 1.5 | 4 |

**Figure S1:**
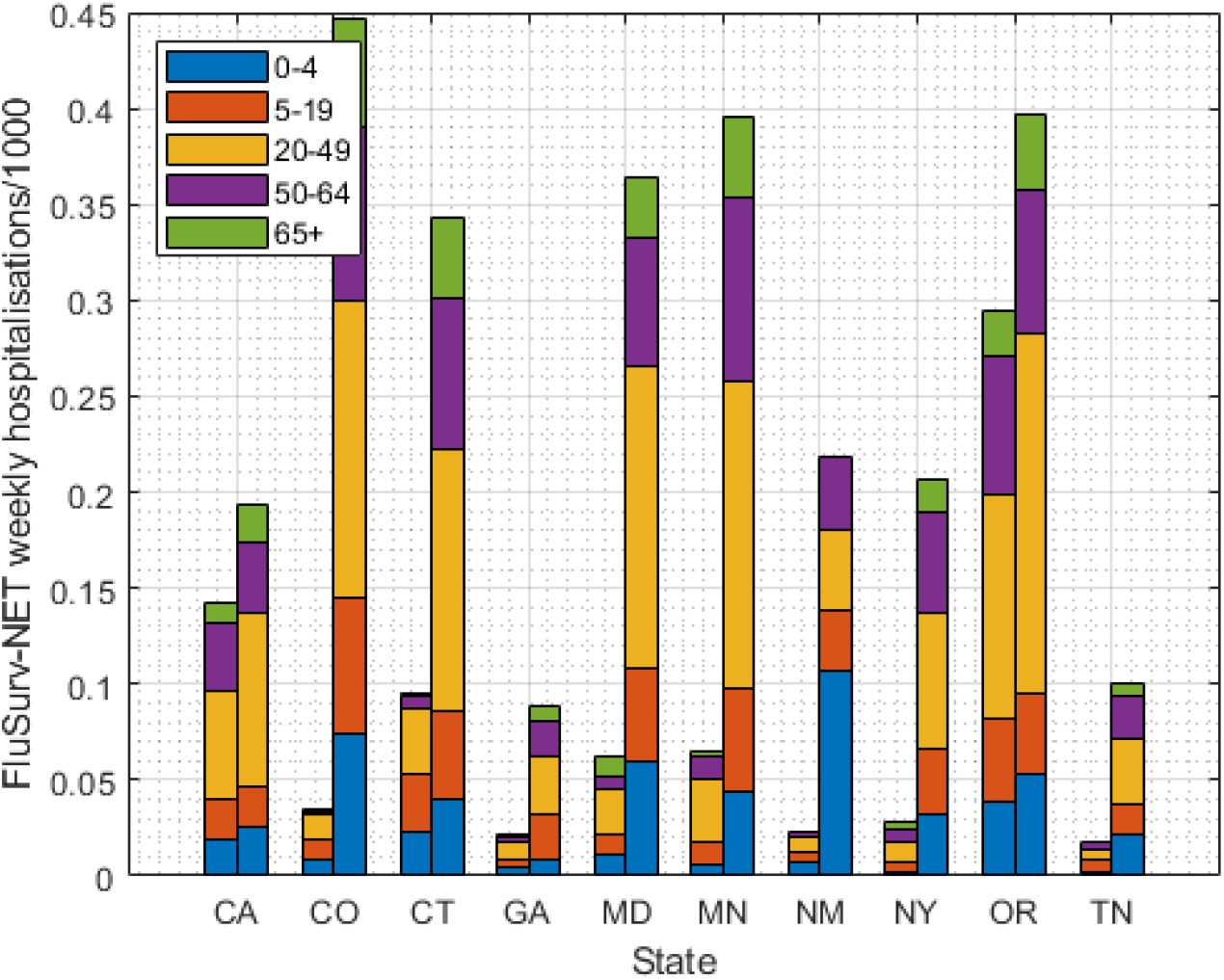
Cumulative hospitalisations by state in the first (left) and second (right) waves of the 2009 influenza pandemic. The second wave is counted from week 35(inclusive), the week of 1st September.

**Figure S2:**
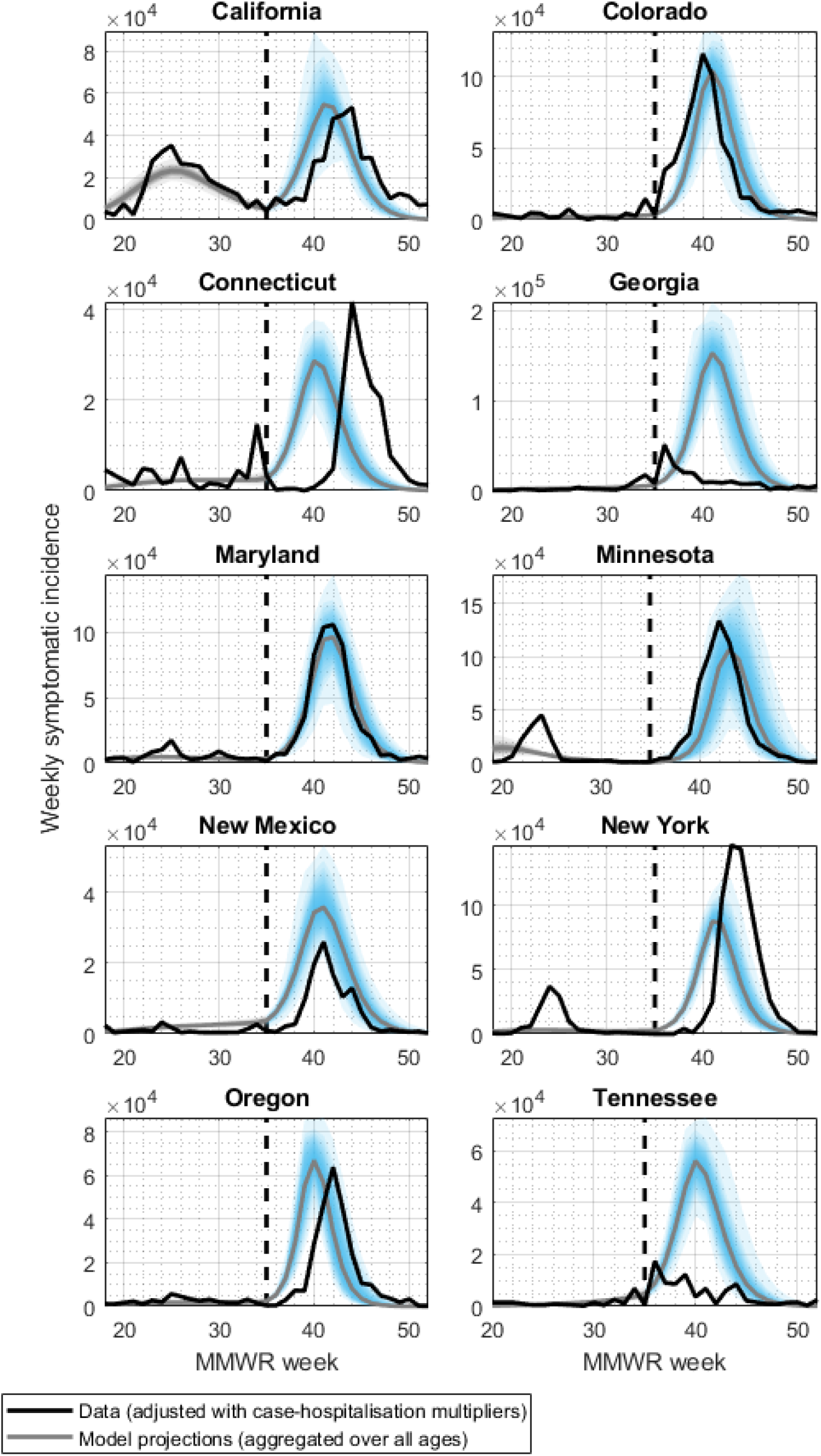
Model calibrations and projections by state.

**Figure S3:**
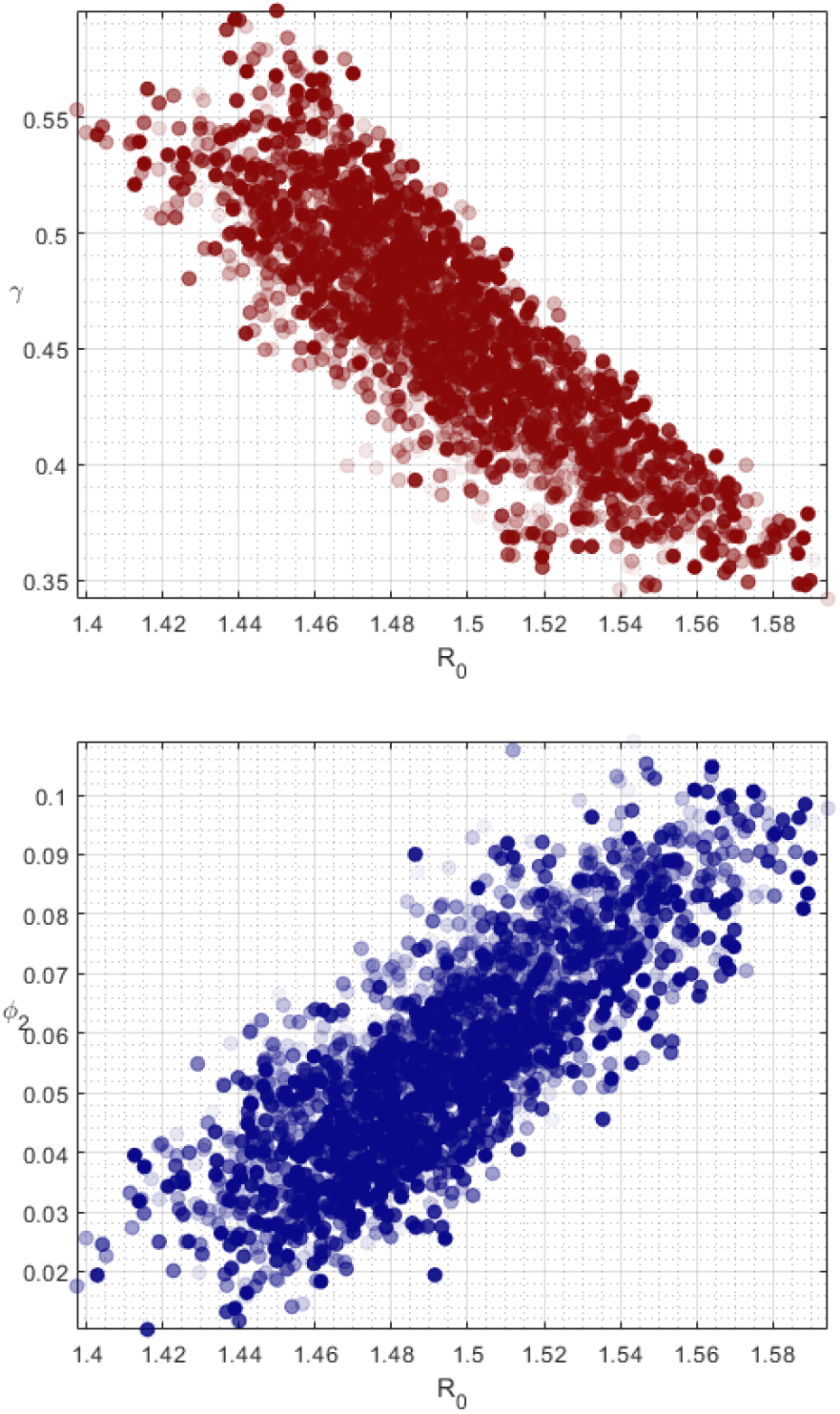
Marginal densities of the Bayesian fit to first-wave data for California: basic reproductive number *R*_0_ and recovery rate *γ* (top); *R*_0_ and amplitude of seasonality *φ*_2_ (bottom).

**Figure S4:**
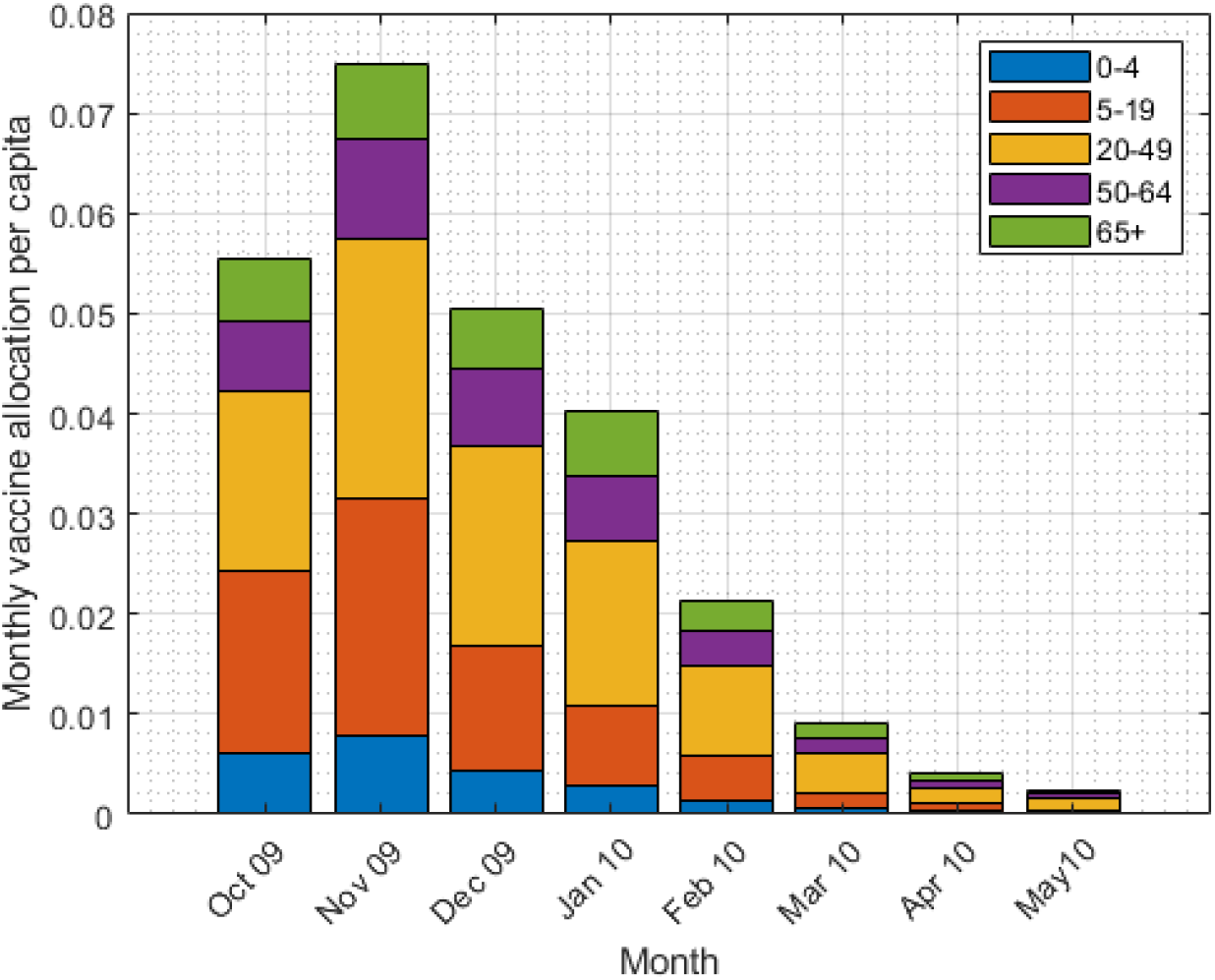
2009 influenza pandemic vaccine roll-out in the USA.

## References

[1] Viggo Andreasen, Cécile Viboud, and Lone Simonsen. Epidemiologic Characterization of the 1918 Influenza Pandemic Summer Wave in Copenhagen: Implications for Pandemic Control Strategies. The Journal of Infectious Diseases, 197(2):270–278, 01 2008.

[2] Guillaume Béraud, Sabine Kazmercziak, Philippe Beutels, Daniel Levy-Bruhl, Xavier Lenne, Nathalie Mielcarek, Yazdan Yazdanpanah, Pierre-Yves Boëlle, Niel Hens, and Benoit Dervaux. The french connection: The first large population-based contact survey in france relevant for the spread of infectious diseases. PLOS ONE, 10(7):1–22, 07 2015.

[3] Yang Cao, Ayako Hiyoshi, and Scott Montgomery. Covid-19 case-fatality rate and demographic and socioeconomic influencers: worldwide spatial regression analysis based on country-level data. BMJ Open, 10(11), 2020.

[4] Dennis L. Chao, M. Elizabeth Halloran, and Jr Longini, Ira M. School opening dates predict pandemic influenza A(H1N1) outbreaks in the United States. The Journal of Infectious Diseases, 202(6):877–880, 09 2010.

[5] Simon de Lusignan, Ray Borrow, Manasa Tripathy, Ezra Linley, Maria Zambon, Katja Hoschler, Filipa Ferreira, Nick Andrews, Ivelina Yonova, Mariya Hriskova, Imran Rafi, and Richard Pebody. Serological surveillance of influenza in an english sentinel network: pilot study protocol. BMJ Open, 9(3), 2019.

[6] Ilaria Dorigatti, Simon Cauchemez, and Neil M. Ferguson. Increased transmissibility explains the third wave of infection by the 2009 h1n1 pandemic virus in england. Proceedings of the National Academy of Sciences, 110(33):13422–13427, 2013.

[7] Anne Ewing, Elizabeth Lee, Cécile Viboud, and Shweta Bansal. Contact, travel, and transmission: The impact of winter holidays on influenza dynamics in the united states. Journal of Infectious Diseases, 215:jiw642. 12 2016.

[8] Anne Ewing, Elizabeth Lee, Cécile Viboud, and Shweta Bansal. Contact, travel, and transmission: The impact of winter holidays on influenza dynamics in the united states. Journal of Infectious Diseases, 215:jiw642. 12 2016.

[9] Anne Ewing, Elizabeth Lee, Cécile Viboud, and Shweta Bansal. Contact, travel, and transmission: The impact of winter holidays on influenza dynamics in the united states. Journal of Infectious Diseases, 215:jiw642. 12 2016.

[10] Harvey V. Fineberg. Pandemic preparedness and response — lessons from the h1n1 influenza of 2009. New England Journal of Medicine, 370(14):1335–1342, 2014. PMID: 24693893.

[11] Heikki Haario, Eero Saksman, and Johanna Tamminen. An adaptive metropolis algorithm. Bernoulli, 7(2):223–242, 04 2001.

[12] Charlotte Jackson, Emilia Vynnycky, and Punam Mangtani. The Relationship Between School Holidays and Transmission of Influenza in England and Wales. American Journal of Epidemiology, 184(9):644–651, 11 2016.

[13] A Kucharski, H Mills, A Pinsent, C Fraser, M Van Kerkhove, CA Donnelly, and S Riley. Distinguishing between reservoir exposure and human-to-human transmission for emerging pathogens using case onset data. PLOS Currents Outbreaks, 2014.

[14] Michael J Mina, C Jessica E Metcalf, Adrian B McDermott, Daniel C Douek, Jeremy Farrar, and Bryan T Grenfell. Science forum: A global lmmunological observatory to meet a time of pandemics. eLife, 9:e58989, jun 2020.

[15] Joël Mossong, Niel Hens, Mark Jit, Philippe Beutels, Kari Auranen, Rafael Mikolajczyk, Marco Massari, Stefania Salmaso, Gianpaolo Scalia Tomba, Jacco Wallinga, Janneke Heijne, Malgorzata Sadkowska-Todys, Magdalena Rosinska, and W. John Edmunds. Social contacts and mixing patterns relevant to the spread of infectious diseases. PLOS Medicine, 5(3):1–1, 03 2008.

[16] O T Mytton, P D Rutter, and L J Donaldson. Influenza a(h1n1)pdm09 in england, 2009 to 2011: a greater burden of severe illness in the year after the pandemic than in the pandemic year. Eurosurveillance, 17(14), 2012.

[17] Carrie Reed, Frederick J. Angulo, David L. Swerdlow, Marc Lipsitch, Martin I. Meltzer, Daniel B. Jernigan, and Lyn Finelli. Science forum: Viral factors in influenza pandemic risk assessment. Emerging Infectious Diseases, 15:e18491, 2009.

[18] Carrie Reed, Sandra S. Chaves, Pam Daily Kirley, Ruth Emerson, Deborah Aragon, Emily B. Hancock, Lisa Butler, Joan Baumbach, Gary Hollick, Nancy M. Bennett, Matthew R. Laidler, Ann Thomas, Martin I. Meltzer, and Lyn Finelli. Estimating influenza disease burden from population-based surveillance data in the united states. PLOS ONE, 10(3):1–13, 03 2015.

[19] Jeffrey Shaman, Virginia E. Pitzer, Cécile Viboud, Bryan T. Grenfell, and Marc Lipsitch. Absolute humidity and the seasonal onset of influenza in the continental united states. PLOS Biology, 8(2):1–13, 02 2010.

[20] Sundar S. Shrestha, David L. Swerdlow, Rebekah H. Borse, Vimalanand S. Prabhu, Lyn Finelli, Charisma Y. Atkins, Kwame Owusu-Edusei, Beth Bell, Paul S. Mead, Matthew Biggerstaff, Lynnette Brammer, Heidi Davidson, Daniel Jernigan, Michael A. Jhung, Laurie A. Kamimoto, Toby L. Merlin, Mackenzie Nowell, Stephen C. Redd, Carrie Reed, Anne Schuchat, and Martin I. Meltzer. Estimating the burden of 2009 pandemic influenza a (h1n1) in the united states (april 2009–april 2010). Clinical Infectious Diseases, 52:S75–S82, 01 2011.

[21] Jonas Vlachos, Edvin Hertegård, and Helena B. Svaleryd. The effects of school closures on sars-cov-2 among parents and teachers. Proceedings of the National Academy of Sciences, 118(9), 2021.

